# Collaborative research methods and best practice with children and young people: Protocol for a mixed-method review of the health and social sciences literature

**DOI:** 10.1101/2022.01.28.22270026

**Authors:** R. Nowland, L. Robertson, N. Farrelly, A. Roy, D. Sharpe, C. Harris, N. Morocza, C. Larkins

## Abstract

**Introduction:** Children and young people have the right to participate in research on matters that affect them, and their contribution improves research quality and insights from findings. Discrete participatory approaches are used across different disciplines. This review will provide a synthesis of existing literature from different disciplines by working with young people and adults experienced in participatory research to develop a broad definition of child and youth led research and to identify best practice.

**Methods and Analysis:** Comprehensive searches will be conducted in eight electronic databases (PsycINFO, Medline, CINAHL, Embase, SocINDEX, ASSIA: Applied Social Sciences Index and Abstracts (Proquest), Social Care Online and SCOPUS). Grey literature reports will also be sourced using Google searching. Eligible studies will be English language primary studies and reviews on collaborative research with children and young people (aged 5-25 years). Qualitative and quantitative data will be integrated in a single qualitative synthesis following the JBI convergent integrated approach. Study quality will be assessed by developed checklists based on existing participation tools co-created with the project steering group and co-creation activities with young people.

**Ethics and Dissemination:** Ethical approval is not required as no primary data will be collected. The review will develop guidance on best practice for collaborative research with children and young people, synthesising learnings from a wide variety of disciplines. Dissemination will be via peer-reviewed publications, presentations at academic conferences and lay summaries for various stakeholders. Opportunities for co-creation of outputs will be sought with the young researchers and the project steering committee.

PROSPERO registration number: CRD42021246378

**Article Summary:** *Strengths and limitations of the study:*

- First systematic review to synthesise findings across different approaches to collaborative research with children and young people
- Research focus, questions and analysis framework have been co-designed with young researchers experienced in participatory research
- Mixed method review methodology will enable an in-depth evidence synthesis across a disparate evidence base
- Provides an interdisciplinary perspective on best practice for collaborative research with children and young people

## Introduction

It is widely acknowledged that children and young people have the right to participate in research on matters that affect their lives, and that their contribution to research adds value to the research processes and outcomes. Involving children and young people as partners in the research process improves research design and refines research priorities, increases the accessibility and attractiveness of research methods and ensures that children and young people’s perspectives are represented in analysis and outputs providing fresh insights and recommendations based on their lived experience.^1^ The right to participate in research is implicit in the 1989 United Nation’s Convention on the Rights of the Child.^2^ It is articulated explicitly in the 2012 Council of Europe Recommendation on Children’s Participation^3^ which notes that member states (including the UK) should:

> *stimulate research on, with and by children and young people, with a view to enabling better understanding of the views and experiences of children and young people, identifying obstacles to their participation and ways of overcoming them* (^3^p9)

The paradigm shift from ‘research on’ to ‘research with and by children and young people’ is of particular significance here as it covers approaches to health and social research that may be called ‘participatory’, in which children and young people take a greater or lesser lead in empirical studies. Increasingly research funders (e.g. Economic Social Research Council, National Institute for Health Research (NIHR)) are expecting children and young people to be research advisors and/or co-researchers, with statements of patient and public involvement being required in funding applications. For example, NIHR in their UK ten year plan for patient and public involvement and engagement published in 2015 commit to having “a population actively involved in research to improve health and wellbeing for themselves, their family and their communities” and the “public as partners in everything we do”.^4^ Since 2012, a number of systematic or mapping reviews have been conducted on participatory research, however apart from reviews by Rouncefield-Swales^1^ and Wilson et al.^5^ - which focus on health research - there has not been a synthesis involving different disciplines on participatory research in which children or young people collaborate with adult researchers and/or take a lead in particular aspects of the research. This review co-produced with young people and adults experienced in participatory research, develops a broad definition of collaborative research with children and young people (i.e. children and/or young people explicitly involved in at least one stage of the research process beyond just generating data and involvement in dissemination or recruitment of participants). It draws on learning from different disciplines/approaches, including Youth Participatory Action Research (YPAR), public and patient engagement, citizen science, community-based peer research and some forms of collaborative research with children and young people.

As mentioned, involving children and young people as collaborators in the research process not only impacts on research design and quality but it can also produce creative and situated forms of ‘learning in action’ (^6^p359) as well as ‘reflexive processes of social engagement’ (^6^p359), which create new spaces for generating and using knowledge.^6^ However, achieving these potential benefits is known to be challenging as it can be hard to ensure that power is distributed, that children and young people’s perspectives are valued, and that research is clearly linked into effective strategies for achieving personal and social change.^7-10^ There continues to be a need for more guidance, particularly on collaborating with marginalised children and young people in ways that enables them to genuinely lead.^11^

In addition to being left out of knowledge production in the ways that adults experience (due to the intersections of ‘race’, ethnicity, gender, class, sexuality and disability), children and young people who experience discrimination through intersecting social ontologies, social categories and social relations face further marginalisation in research.^12^ The exclusion of children and young people is pervasive due to dominant conceptions of children and young people as an homogenised social category represented as incompetent, vulnerable, politically immature and needing the completion of education in order to deserve recognition as citizens and as competent researchers.^13^ Young people are often conceived of as apathetic or troublemakers, rather than recognising how young people are alienated by neo-liberal practices .^14^ The battle over what counts as evidence^15^ can also render children and young people’s perspectives and sometimes their chosen means of expression, less valid than scientific orthodoxy.

Where children and young people are included in research, they may be provided with information, or experience being ‘researched on’, however their influence over the priorities to investigate, and changes in guidance and practice is poorly evidenced. In 1999, Pole et al^16^ noted that, despite the turn towards participatory methods across multiple disciplines, children and young people do not have enough research capital to make them serious stakeholders in the research process. Brownlie^7^ repeated this, echoing the concern that “children and young people remain a long way from the emancipatory call of ‘nothing about us, without us’”(^7^p711). And still, a decade on, Lohmeyer^8^ repeats that “In theory, youth participatory methods are participant-led, and adults are involved in the process. However, there are social, historical, procedural and institutional barriers that make this ideal all but unachievable”(^8^p44). This is despite the fact that some young people are “keen as f**k” to participate.^8^

Unless these barriers to collaborative research with children and young people are fully understood and strategies for overcoming the challenges are shared, research risks being perpetuated as yet another form of symbolic violence.^17 18^ That is, it will create conditions which perpetuate and normalise children and young people’s subordinate position in processes of knowledge creation. Or, peer-led research may become a mechanism through which children and young people are exploited as lower paid or unpaid labour, to access young communities who are suspicious of mainstream health and social science research without allowing them power to identify what issues need investigating. There is therefore need for greater attention to the precise mechanisms, methods and reflexive stances which enable children and young people to lead research.^18^

Questions remain, however, about the kind of knowledge that is generated by collaborative research methods and attention to what we mean by concepts such as knowledge and epistemology. Young researchers cocreate methods, including digital methods^19^, photo-walks^20^, map-making^21^ and storytelling^22^ which extend beyond traditional methods. Young researchers highlight that these methods are experienced positively by research participants^23,24^ and hence these co-creative approaches acknowledge shared responsibilities and skills in health and social research.^25^ But these methods are not always valued by end users of research outputs, resulting in biases towards research that is not always congruent with children and young people’s interests, concerns and contexts. Policy actors, funders and commissioners may need greater awareness of a diversity of approaches to rigour, quality and impact^26^, and may need to extend their understanding of health and social research to also recognise the validity that arises from greater degrees of participation.^27^ Evidence that can demonstrate how the knowledge from collaborative research with children and young people can be valued by and acted on by decision-makers may therefore provide further benefits.

What is needed is a synthesis of epistemologies and methodologies across a broad range of different disciplines to establish key contexts for successful research by and with children and young people. The current review addresses this gap by establishing precise mechanisms, methods and reflexive stances which enable children and young people to lead and collaborate as partners in research identifying best practice from existing evidence. The review will inform both researchers and policy actors, funders and commissioners of the diversity of approaches that may be appropriate to enable collaborative research with children and young people whilst maintaining academic rigour and quality. Barriers and challenges will be highlighted to ensure power imbalances are addressed and ways of working with marginalised groups will be identified. The review will be useful to guide future collaborative research with children and young people but will also identify key gaps in the evidence base where future work needs to be conducted.

### Aim of the review

To identify theoretical principles and practice modes and mechanisms of effective collaborative research with children and young people in the field of health and social sciences, that are generalisable as a basis for designing effective peer research projects, protocols and establishing best practice.

The mixed methods review will scope and synthesise existing knowledge about best practice in conducting collaborative research with children and young people using the following research questions co-created with young people and adults experienced in participatory research:

1. What are the opportunities, barriers and tensions in youth peer research and how can these be understood and addressed?
2. What are the different modes and mechanisms of doing peer research? Which of these are valued, by whom, in which contexts and why?
3. How is success, impact and change documented, understood, negotiated and evaluated in peer research?

Question 1 focusses specifically on issues identified by young people experienced in participatory research as critical involving *cushions* (e.g. negotiated support with tasks, skills, decision making and managing the emotional impact of conducting research), *credibility, collaboration* and *change*. More detailed sub-questions have been devised to address these highlighted issues:

a. How do young and adult researchers ensure that young researchers have the *cushions* they want throughout the research process?
b. Which processes and structures ensure *collaborative* peer research is acceptable and accessible to the diversity of children and young people (age, identity, experience of discrimination, economic situations)?
c. How can we ensure that peer research is maximised in terms of strengthening claims to knowledge and *credibility*, conveying convincing stories, linking to current opportunities and minimising risk of negative attention?
d. Which processes and/or structures help ensure productive relationships between stakeholders, allies, contexts and resources to support the use of evidence to make *change* possible?

Ethics, safety, inclusion and power as themes relating to peer research will be considered across all research questions. We will also report on the topics into which peer research has been conducted and examine differences in modes, mechanisms and success across different topics.

## Methods and Analysis

This protocol is guided by the Preferred Items for Reporting of Systematic Reviews and Meta-Analyses Protocols (PRISMA) checklist^28^ (online supplemental appendix 1) and the Joanna Briggs Institute (JBI) methodology for mixed-methods systematic reviews.^29^

### Study registration

Based on the PRISMA guidelines^28^, the protocol for this systematic review was registered on the international database of prospectively registered systematic reviews in health and social care, PROSPERO. Any important protocol amendments will be recorded in PROSPERO and published with the results of the review.

The mixed methods review will involve: 1) a systematic review of reviews on peer research and 2) realist synthesis of process and descriptive papers defined as those describing lessons learnt or a description of the programme, process or training of a peer model and articles that focused on the peers themselves and their experiences within a peer model/approach (distinction from Vaughn et al.^30^).

The systematic review of reviews will identify and establish the core models and methods used in collaborative research with children and young people and the realist synthesis will offer a more nuanced understanding of what works in peer research, for whom, in what contexts and why. Findings will be triangulated and used to develop a critical appraisal tool to assess peer research.

### Eligibility criteria

Studies and reviews will be selected according to the criteria set out below:

#### Types of studies

We will include systematic and scoping reviews, descriptive and process papers (using the distinction made by Vaughn et al.^30^) relating to peer research, including also grey literature reviews/reports. We will exclude papers that are exclusively empirical papers without description of process or reflections, dissertations, editorials, opinion pieces, commentaries, book or movie reviews, protocols, reports, case studies and erratum. We will only include studies about peer research with children and young people. We will exclude studies examining peer research in adult populations. Only studies written in English will be included and those published after 2000 (due to the expediential growth in young people’s involvement in social research from 2000 onwards).

We will use a wide definition of peer research and include all reviews and process/descriptive papers including a wide range of terms used to describe peer research (i.e. participatory research, YPAR, community-led research, peer research informed social action etc.).

#### Participants

We will include peer research involving children and young people (aged 5-25 years) and exclude peer research conducted with adults. Articles about research with primary school aged children will be included to extrapolate potentially generalisable findings on peer research to an older population of children and young people, but we will be mindful of differences in developmental stages and needs.

Peer research (including community-based participatory research, peer led research, youth inquiry, co-production, citizen science, youth participatory action research, peer research informed social action).

#### Outcomes

Theoretical principles, practice and mechanisms and findings in relation to power, inclusivity, ethics, safeguarding, learning, methods, and impact.

We report on other important or critical factors and influencers of best practice in peer research highlighted by selected papers.

### Search strategy

We recruited a review steering group involving participants from Youth Endowment Fund, study partners, appointed advisors and experienced young researchers from marginalised groups, academics experienced in youth participation and relevant third sector professionals and policy actors. Online discussions with this group (n = 18) were held in the form of a week-long civic hackathon (^31^; creative problem-solving sessions conducted once a day (1 ½ hours long) for a full week in March 2021) involving activities to enable:

- Reflection and sharing of ideas about key concepts and challenges in peer research
- Reflection and definition of a proportionate systematic approach and relevant inclusion criteria
- Agreement of research questions, inquiry themes and focus for the review

The findings from the online hackathon informed the focus of the research, search strategy, inclusion and exclusion criteria and framework for synthesis.

In addition, we conducted a priori scoping searches to identify key review papers in this specific research area which also informed our search strategy.

We will use the Preferred Reporting Items for Systematic Review and Meta-Analysis checklist (PRISMA;^32^) as a framework for the review.

We plan to conduct searches on eight bibliographic databases:

> PsycINFO, Medline, CINAHL, Embase, SocINDEX, ASSIA: Applied Social Sciences Index and Abstracts (Proquest), Social Care Online and SCOPUS

Handsearching will also be used, involving forward and backward chaining and examination of references lists from reviews and key papers in this research area. We will also check author’s personal files for any key studies. In accordance with PRISMA guidelines^32^, the number of search results will be recorded at each stage of the study identification process. In order to locate wider reviews on peer research that have been conducted we will include grey literature reports, which will be obtained through Google searching using the key words (first 200 hits will be screened).

The following search terms have been developed following a priori scoping exercises and online forum exercises with experienced young peer researchers and stakeholders:

> (Child/ or Adolescent/ or child or children or kid or kids or girl* or boy* or adolescen* or teen* or Youth* young people or young adult or young person or young men or young women)

AND

> Community-based participatory research/ or participatory research* or participatory method* or participatory approach* or participatory design or participatory model* or user led research or peer led research or peer research* or consumer led research or action research or youth inquir* or co-produc* or coproduc* or co-research or coresearch or co-creation or cocreation or co-design* or codesign* or co-develop* or codevelop* or co-investigator* or coinvestigator* or citizen science or citizen scientist or YPAR or advisory group* or advisory council or youth participation or young involved or child led research* or peer model or research partner or social action)

The search strategy will be adapted to meet the truncation and Boolean operations of each database as appropriate. An example of the search strategy used for Medline database is presented (online supplemental appendix 2).

### Study Selection

Papers identified from database searches will be downloaded to Endnote and any duplicates removed. Screening by title and abstract will be conducted in Rayyan independently by one of the authors, with at least 20% of the papers screened by another author. Decisions will be based on the inclusion and exclusion criteria.

Once screening by title and abstract is complete, papers selected for full text screening will be sourced and then examined by one author independently, with at least 20% of the papers screened by another author. Reasons for exclusion will be noted at this stage.

Agreement at all stages will be made by consensus, and any disagreements regarding inclusion will be discussed with a third reviewer. Inter-rater reliability will be recorded at each screening stage (i.e. title, abstract and full text screening).

### Data extraction

Following screening, data will be extracted from all selected texts using a data extraction sheet with a framework developed and co-created with the steering group. As suggested by Daudt, van Mossel, and Scott^33^ (2013) at least 20% of data extracted will be charted by two authors independently using the data extraction tool. Once sufficient agreement (> 80%) has been reached in the test phase, authors will apply the tool to the remaining studies. Disagreements between the authors completing the data extraction will be resolved through discussion, including the involvement of a third reviewer where necessary. It is expected that data extraction will include key study characteristics, participant characteristics, definitions of peer research, models and mechanisms (focussing specifically on research approaches and processes identified in hackathon activities) relationships, attitudes, approaches, resources, distribution of leadership, timescales, and change) and co-created frameworks based on identified challenges and tensions in peer research centred on *cushions, credibility, collaboration* and *change*. We will also chart any other important or critical factors and influencers of best practice in peer research highlighted within selected papers.

During the data extraction stage, the research team will meet on a regular basis to discuss progress, and to consider decisions regarding the relevance and adequacy of the data collection tool. Those discussions will be documented along with any changes to the study protocol and data extraction. Study authors will be contacted if additional information is required (e.g. context related details of the study).

### Assessment of Methodological Quality

Two authors will independently assess the research quality and bias of each of the included articles involving studies of peer research using developed checklists based on existing participation tools (e.g. Larkins et al.’s Participation Lattice^34^; Shier’s analytical tool^35^) co-created with the steering group and based on the results of activities in the hackathon. Discrepancies between the review authors will be resolved by discussion, consulting a third review author where necessary.

Two authors will independently assess the research quality and bias of all the review studies on peer research included using the AMSTAR 2 Appraisal Tool^36^ for systematic reviews. Inter-rater reliability will be reported and any discrepancies between authors will be resolved through discussion or where necessary a third author will be consulted.

### Data synthesis

Data extracted will be collated, summarised and synthesised narratively. Data will be presented as tables, charts and/or visual maps in an aggregate rather than individual basis, to provide an overview of the research field, summarise findings, identify gaps in the literature and make recommendations for future research. Data analysis will be conducted in two phases: 1) narrative synthesis of theoretical principles (i.e. definitions of peer research) and mechanisms/methods used and 2) analysis of findings around the co-produced thematic framework *cushions, credibility, collaboration* and *change* and 3) content and thematic analysis using a co-created approach realist framework. We will explore youth characteristics and contextual factors that influence what works for peer research with young people.

### Patient and Public Involvement statement

The public were involved from the very start of developing the protocol. Young researchers and non-academic third sector professionals (service providers and funders) took part in a series of online discussions with academics. This was framed as a civic hackathon^31^, that is a series of online events held in quick succession, with the aim of identifying what is currently understood by the term peer research by and with children and young people, to explore the challenges and potential of these approaches and to create a set of questions to guide the review. Four online events were conducted, of around 90 minutes each, to frame the review. The events were facilitated by senior academics experienced in participatory research with young people. We used visual aids and online scribing to elicit the perspectives of young people and adults experienced in participatory research and then guest academics were asked to respond to this. At the end of every meeting we created a 3 minute summary of key discussion points and perspectives and shared this, along with the visual and text notes of the meeting, to support the participation of those who could not attend on specific days. Contributors to these non-synchronous discussions tended to be academics. At the start of every meeting we reviewed the story of our discussions so far, and summarised content that had been provided in between meetings. At the end of the third meeting, ideas generated to date were used to draft initial questions for the review. These were amended and finalised at the fourth meeting.

Whilst the review has been underway a further two online events have been held to discuss emerging findings and potential outputs and a further four events are planned to enable young researchers to contribute to at least one accessible output (an audio podcast has been planned) and all academic articles. Young people have decided that the podcast will be shared on an open access platform codesigned by young researchers for young researchers. All participants in these activities either contributed as part of paid roles or received a thank you in the form of vouchers.

## Supporting information

PRISMA checklist

## Data Availability

All data produced in the present work are contained in the manuscript

## Ethics and Dissemination

Ethical approval and consent to participate are not required for the proposed systematic review as no primary data will be collected. The findings of the mixed methods review will be written up as a report which will directly inform peer research training for the Peer Research and Social Action Network, funded by the Youth Endowment Fund together with the #iwill Fund and the Co-op Foundation. The Peer Research and Social Action Network will support young people affected by violence to become Peer Researchers and Changemakers. We will also explore opportunities with youth peer researchers to co-create accessible outputs to be disseminated through peer research networks. We expect that the findings will be written up in peer reviewed academic journals as a systematic review of reviews, realist synthesis reviews of papers about processes of peer research, and intergenerational reflections on the review process.

## Authors Contributions

CL is the guarantor. RN and CL drafted the manuscript. All authors contributed to the development of the selection criteria, the risk of bias assessment strategy and data extraction criteria. RN, CL and CH developed the search strategy. CL, LR, DS, and AR provided expertise on peer research with children and young people. RN and LR provided expertise on systematic review methodology. All authors read, provided feedback and approved the final manuscript.

## Data statement

Datasets (i.e. data extraction charts and analyses) will be available from the University of Central Lancashire’s online repository (CLOK).

## Acknowledgements

The authors thank the project steering committee, youth and adult researchers experienced in peer research that inputted into the development of the research plan and protocol.

